# Measuring vaccine efficacy against infection and disease in clinical trials: sources and magnitude of bias in COVID-19 vaccine efficacy estimates

**DOI:** 10.1101/2021.07.30.21260912

**Authors:** Lucy R. Williams, Neil M. Ferguson, Christl A. Donnelly, Nicholas C. Grassly

## Abstract

**Background:** Phase III trials have estimated COVID-19 vaccine efficacy (VE) against symptomatic and asymptomatic infection. We explore the direction and magnitude of potential biases in these estimates and their implications for vaccine protection against infection and against disease in breakthrough infections.

**Methods:** We developed a mathematical model that accounts for natural and vaccine-induced immunity, changes in serostatus and imperfect sensitivity and specificity of tests for infection and antibodies. We estimated expected biases in VE against symptomatic, asymptomatic and any SARS-CoV-2 infections and against disease following infection for a range of vaccine characteristics and measurement approaches, and the likely overall biases for published trial results that included asymptomatic infections.

**Results:** VE against asymptomatic infection measured by PCR or serology is expected to be low or negative for vaccines that prevent disease but not infection. VE against any infection is overestimated when asymptomatic infections are less likely to be detected than symptomatic infections and the vaccine protects against symptom development. A competing bias towards underestimation arises for estimates based on tests with imperfect specificity, especially when testing is performed frequently. Our model indicates considerable uncertainty in Oxford-AstraZeneca ChAdOx1 and Janssen Ad26.COV2.S VE against any infection, with slightly higher than published, bias-adjusted values of 59.0% (95% uncertainty interval [UI] 38.4 to 77.1) and 70.9% (95% UI 49.8 to 80.7) respectively.

**Conclusion:** Multiple biases are likely to influence COVID-19 VE estimates, potentially explaining the observed difference between ChAdOx1 and Ad26.COV2.S vaccines. These biases should be considered when interpreting both efficacy and effectiveness study results.

## Introduction

The coronavirus disease (COVID-19) phase III vaccine trials have demonstrated efficacy against symptomatic infection for multiple vaccine candidates, with estimates ranging from 50% to 95% (1– 3). Yet a vaccine that protects against symptomatic disease may work either by preventing infection, (an infection-blocking vaccine), by preventing the progression to symptoms upon infection (a disease-blocking vaccine), or by a combination of these two mechanisms (Supplementary Figure 1) (4). Understanding the extent to which the COVID-19 vaccines protect against infection is important because the success of each vaccination programme is highly contingent not only on symptomatic cases, but also asymptomatic infection and community transmission (5).

The predominant primary outcome of the COVID-19 vaccine trials is vaccine efficacy against the first case of PCR-confirmed symptomatic disease, *VE*_*sym*_. This is measured by PCR-testing trial participants with COVID-19 symptoms, and is sensitive to the clinical case definition (6). As important secondary outcomes, most trials also measure the incidence of asymptomatic infections, using either i) regular swabbing and PCR-testing or ii) serological testing for anti-nucleocapsid antibodies at pre-specified time intervals (Table 1). The latter allows seroconversion after infection to be identified for trials of vaccines based on the spike, but not inactivated vaccines that include the whole virion (although for inactivated vaccines, it may be possible to infer infection via a rise in SARS-CoV-2 antibodies following a period after vaccination). This allows for vaccine efficacy against asymptomatic infection (*VE*_*asym*_) and vaccine efficacy against any infection (*VE*_*in*_) to be estimated.

**Table 1.**
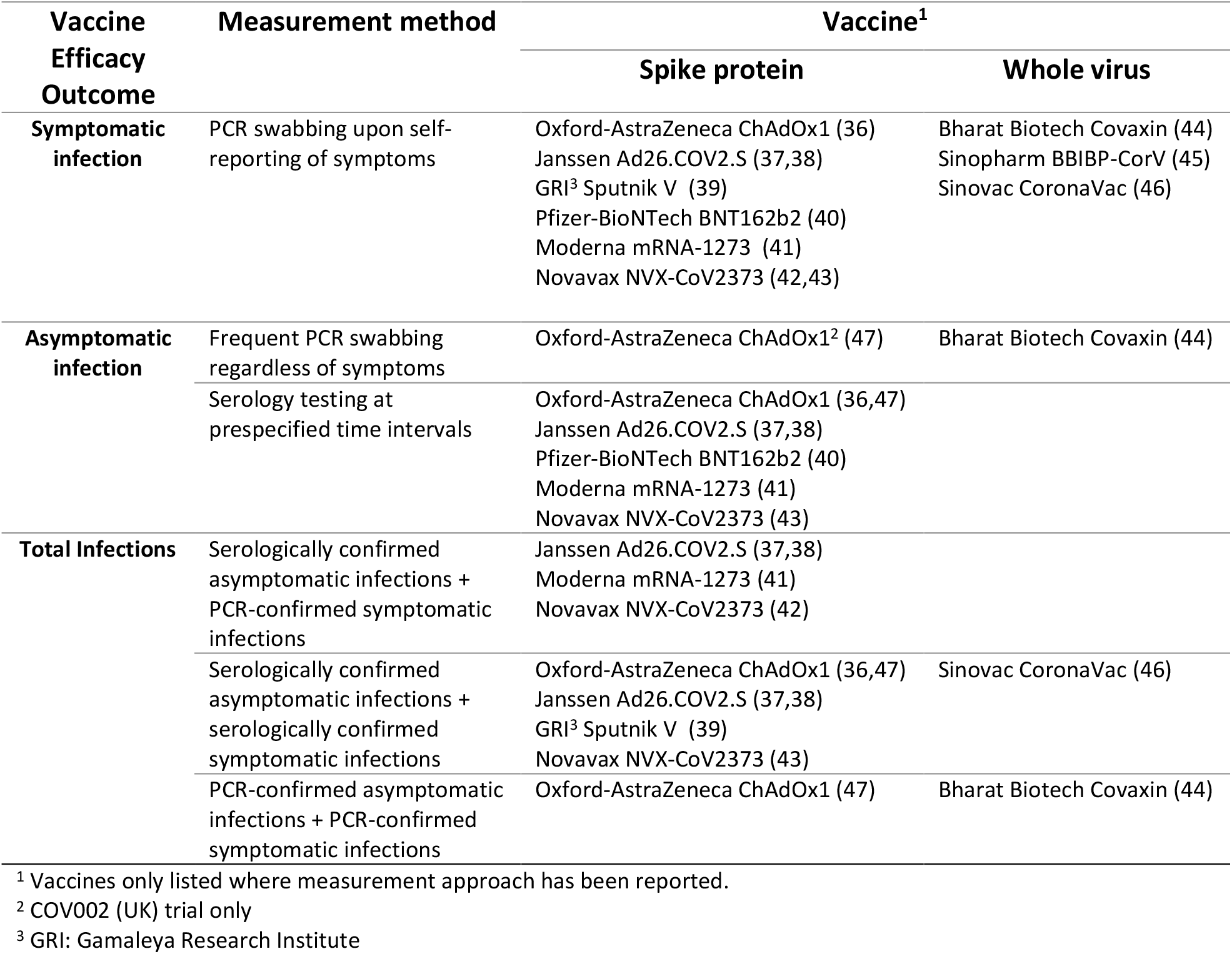
Methods for measuring vaccine efficacy against infection, symptomatic infection and asymptomatic infection in the COVID-19 phase III trials.

*VE*_*asym*_ is a complex outcome due to its relationship with the two mechanisms of vaccine efficacy. A vaccine that protects only against infection will reduce the number of symptomatic and asymptomatic infections in equal proportions, leading to a positive *VE*_*asym*_. Yet a vaccine that protects against progression to symptoms will convert symptomatic cases to asymptomatic, potentially giving a negative *VE*_*asym*_. The counterintuitive interpretation of this outcome has been noted (4,7), but the relationship between *VE*_*asym*_, *VE*_*in*_ and vaccine efficacy against progression to symptoms (*VE*_*pr*_) has not been quantified. To aid the interpretation of *VE*_*asym*_, it is important that the relationship between this outcome and the two main mechanisms of vaccine efficacy is described.

Estimates of vaccine efficacy are known to be biased by factors such as imperfect test sensitivity and specificity, undetected asymptomatic infections, and the accumulation of immunity over time (8–10). However thus far, there has been little discussion on the accuracy and potential biases of the COVID-19 phase III trial estimates (11,12). We developed a conceptual framework and mathematical model of a vaccine trial to investigate the factors affecting observed values of vaccine efficacy. We first illustrate the factors that influence measured *VE*_*asym*_ and derive its relationship with *VE*_*in*_ and *VE*_*pr*_. We then quantify the influence of different biases on vaccine efficacy estimates, notably the impact of i) the build-up of immunity from undetected asymptomatic infections, ii) imperfect test sensitivity and specificity for alternative testing strategies, iii) differential detection of asymptomatic and symptomatic infections, and iv) confounding of vaccine efficacy and probability of symptoms by age. We finish by applying our model to published COVID-19 trial results, to estimate the potential bias in their estimates.

## Methods

### Analytical derivations

Vaccine efficacy (VE) is defined as 1-RR where RR is some measure of the relative risk of the outcome in the vaccine compared with the control arm (13). For most primary and secondary outcomes of the COVID-19 vaccine trials, the relative risk is based on an incidence rate ratio (IRR) such that

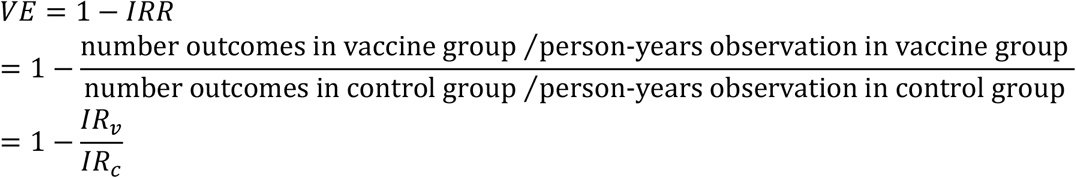

where *IR*_*v*_ and *IR*_*c*_ are the incidence rate in the vaccine and control groups respectively.

For outcomes measured at fixed time points, such as the number of seroconversions to a protein that is not a component of the vaccine, the relative risk can be calculated using the cumulative incidence ratio (CIR) such that

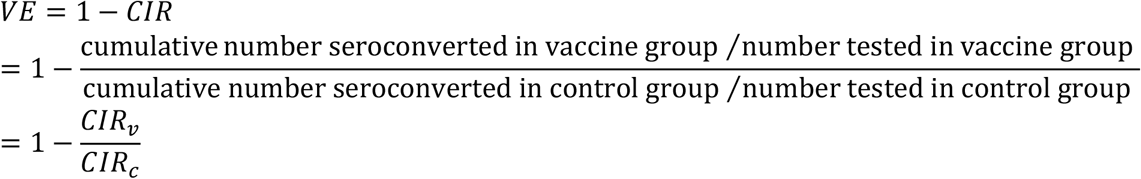

where *CI*_*v*_ and *CI*_*c*_ are the cumulative incidence in the vaccine and control groups respectively. For a partially protective vaccine, vaccine efficacy based on cumulative incidence approximates that based on the incidence rate for low incidence or short follow-up periods, but biases towards zero as follow-up time and incidence increase (10).

For vaccines that protect against infection and/or progression to symptoms, vaccine efficacy against symptomatic infection is given by

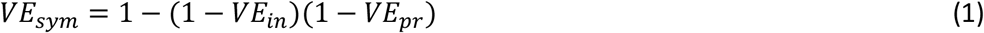

(14). Vaccine efficacy against asymptomatic infection depends on the incidence of asymptomatic infections that are not prevented by the vaccine and on infections that would have become symptomatic, but the vaccine prevents from progressing, such that

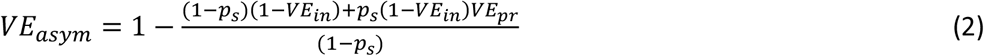

where *p*_*s*_ is the probability of symptom development in the absence of vaccination. Substituting equation (1) into (2) allows *VE*_*in*_ to be derived as a simple function of *VE*_*sym*_ and *VE*_*asym*_

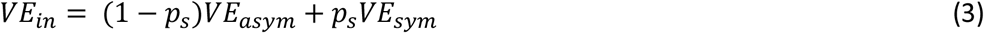

Rearranging equation (1) and substituting equation (3) into equation (1) then provides a solution for *VE*_*pr*_

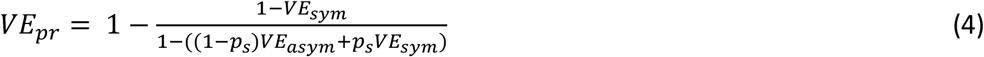

Estimation of confidence intervals for *VEin* and *VEpr* is provided in Supplementary Methods. If asymptomatic infections are less likely to be detected than symptomatic infections, and a vaccine is protective against progression to symptoms (*VE*_*pr*_ > 0), then observed *VE*_*in*_ ≠ true *VE*_*in*_. In this case, the observed *VE*_*in*_ depends on the relative incidence of detected infections, and can be related to the true efficacy by

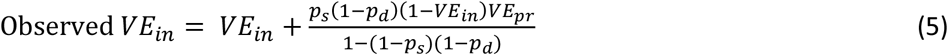

where *p*_*d*_ represents the relative probability of asymptomatic to symptomatic infection detection.

Analytical solutions become more complex when incorporating additional biases, such as those resulting from imperfect test sensitivity and specificity and the acquisition of natural immunity, so we developed a stochastic mathematical (cohort) model of a phase III vaccine trial.

### Mathematical model

The model follows a susceptible, infected, recovered (SIR) structure, implemented as a Markov model, and allows for asymptomatic and symptomatic infections, natural immunity, changes in measured serostatus and imperfect test sensitivity and specificity. The background infection rate, defined as the rate at which a susceptible unvaccinated person becomes infected, is assumed to be constant over time. We assume the “leaky vaccine” model, so for vaccinated participants the background infection rate is reduced by the *VE*_*in*_. The probability of unvaccinated participants developing symptoms upon infection is defined by *p*_*s*_, while for vaccinated participants *p*_*s*_ is reduced by the *VE*_*pr*_. We assume no heterogeneity in population characteristics but perform a sensitivity analysis to assess the effect of variation in *p*_*s*_ and vaccine efficacy by age.

We model two testing approaches for asymptomatic infections, i) weekly PCR testing and ii) serological testing at 1, 2, 6, 12 and 24 months after baseline. We assume that responsive PCR testing detects all symptomatic infections. Observed vaccine efficacy is calculated from the simulated incidence of detected infections in each trial arm. Efficacy is estimated as 1-IRR for all outcomes except those estimated using serology, for which efficacy is estimated as 1-CIR, using the cumulative number of seroconversions detected in each serology assessment up to the present time interval. Point estimates and confidence intervals are given by the mean and 2.5 and 97.5 percentiles of 1000 simulated estimates.

### Application to COVID-19

Applying the model to COVID-19, we assumed a natural probability of developing symptoms upon infection of 0.67 (15), a serology test specificity of 99.84% (16) and sensitivity of 95% and 80% to symptomatic and asymptomatic infections, respectively (17). We used data on the probability of PCR detection over time since infection for individuals without symptoms (18) to estimate the probability of detecting an asymptomatic infection with weekly PCR swabbing (Supplementary Table 1), and we assumed a PCR test specificity of 99.945% (19). We used the model to estimate bias-adjusted vaccine efficacy estimates for two adenovirus vector vaccines that have published trial data on asymptomatic infection, ChAdOx1 (Oxford-AstraZeneca) and Ad26.COV2.S (Janssen), and generated 95% uncertainty intervals (UI) using Latin Hypercube Sampling. We then used rank regression to evaluate the contribution of individual parameters to the biases (Supplementary Methods).

The model is described further in Supplementary Methods and illustrated in Supplementary Figures 2 and 3. The model parameters are provided in Supplementary Table 2.

## Results

### Interpretation of vaccine efficacy against asymptomatic infection

Observed *VE*_*asym*_ was dependent on *VE*_*in*_, *VE*_*pr*_ and the proportion of infections that were symptomatic (Figure 1). For vaccines that worked only by blocking infection, *VE*_*asym*_ was equal to *VE*_*sym*_. For vaccines with efficacy predominantly mediated by prevention of symptom development, *VE*_*asym*_ was low or negative, particularly when a large proportion of infections were naturally symptomatic. *VE*_*in*_ and *VE*_*pr*_ could be estimated from reported values of *VE*_*sym*_ and *VE*_*asym*_ using Equations 3 and 4 (Supplementary Figure 4).

**Figure 1.**
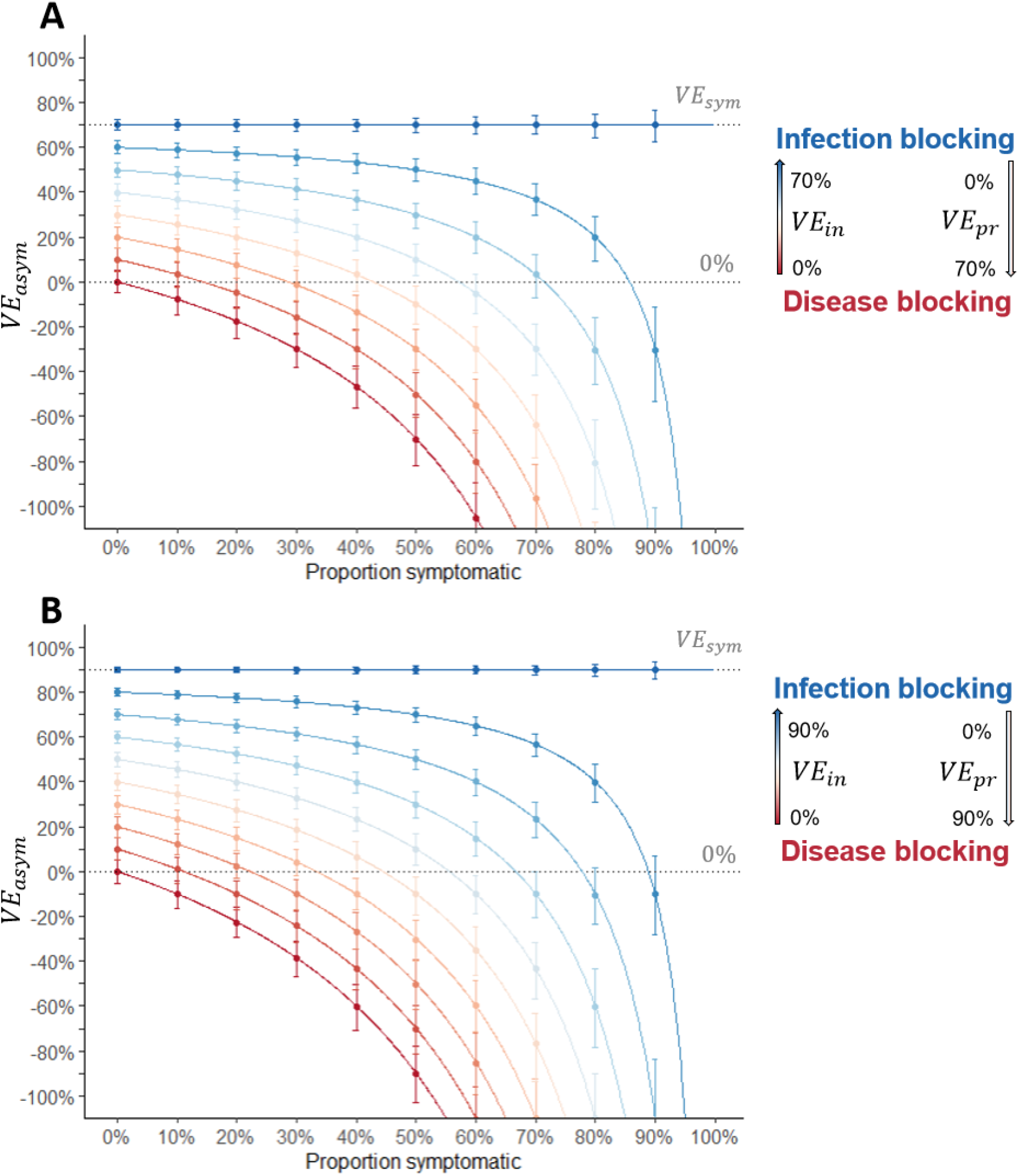
Estimated vaccine efficacy against asymptomatic infection. Red to blue gradient represents the transition from a disease-blocking vaccine (*VE*_*in*_ = 0% and *VE*_*pr*_ = *VE*_*sym*_) to an infection-blocking vaccine (*VE*_*in*_ = *VE*_*sym*_ and *VE*_*pr*_ = 0%). Each successive line represents a 10% increase in *VE*_*in*_, and a corresponding decrease in *VE*_*pr*_ (such that *VE*_*sym*_ remains the same). Lines show predicted values from Equation 2, points and error bars show the observed mean and 2.5 and 97.5 percentiles from 1000 simulations, with efficacy calculated as 1-IRR, censoring after the first infection. A) Vaccine with 70% efficacy against symptomatic infection (e.g., Oxford-AstraZeneca/Janssen vaccines). B) Vaccine with 90% efficacy against symptomatic infection (e.g., Pfizer/Moderna vaccines). *VE*_*in*_= vaccine efficacy against infection, *VE*_*pr*_= vaccine efficacy against progression to symptoms, *VE*_*sym*_= vaccine efficacy against symptomatic infection, IRR = incidence rate ratio.

### Possible biases in the COVID-19 vaccine trials

The build-up of immunity from undetected asymptomatic infections caused *VE*_*sym*_ to bias in opposite directions for infection-blocking and disease-blocking vaccines (Figure 2). For an infection-blocking vaccine, estimated *VE*_*sym*_ decreased over time, with greater decreases observed for higher forces of infection and lower probabilities of symptoms. For a disease-blocking vaccine, a downward bias was only observed when the probability of symptoms was low. Instead, for most combinations of parameters, estimated *VE*_*sym*_ increased slightly over time. Further sensitivity analysis showed that the biases were sensitive to the vaccine efficacy calculation (Supplementary Figure 5).

**Figure 2.**
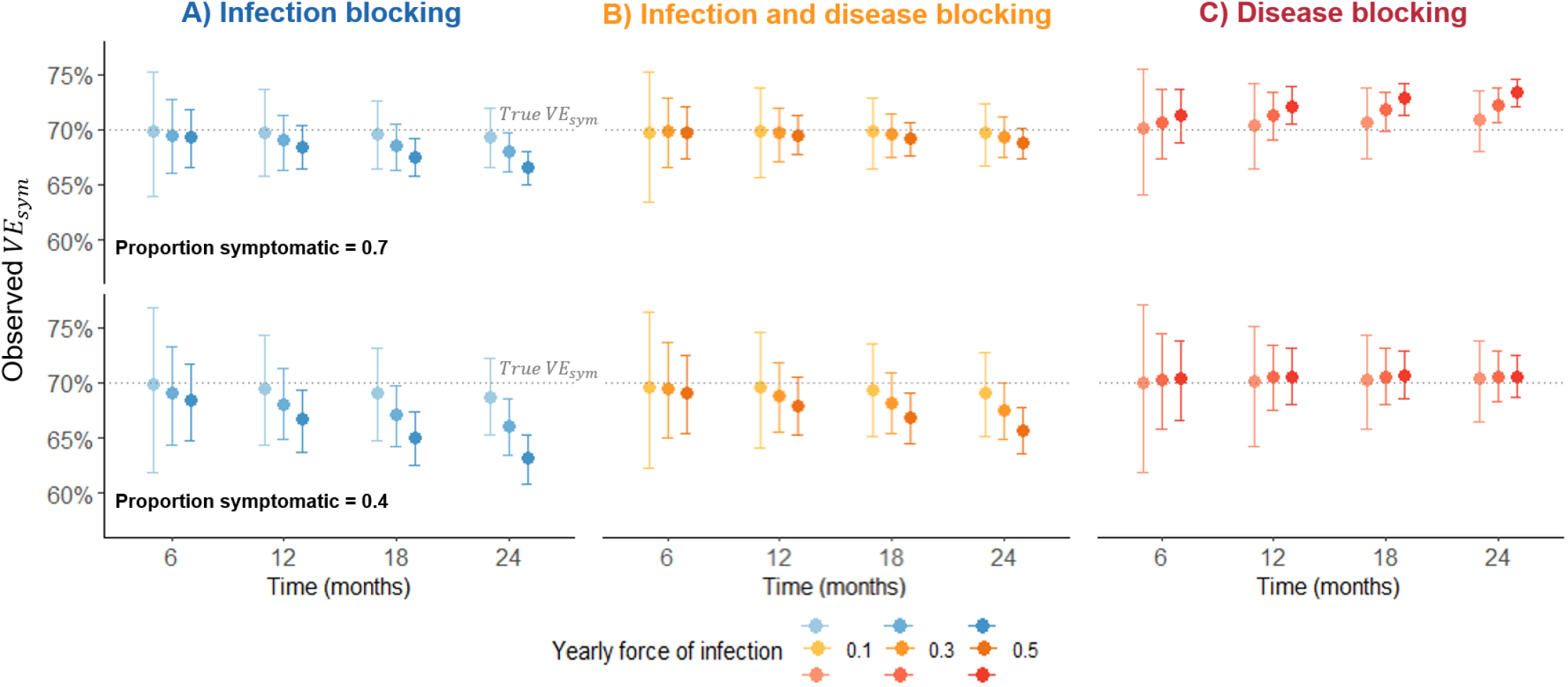
Change in estimated vaccine efficacy against symptomatic infection over a two-year follow-up. Points and error bars show the observed mean and 2.5 and 97.5 percentiles from 1000 simulations, with efficacy calculated as 1-IRR, censoring after the first symptomatic infection. Time = 0 months represents 2 weeks post second dose. Sensitivity and specificity = 100%. True vaccine efficacies: vaccine efficacy against symptomatic infection (*VE*_*sym*_) = 70%, A) vaccine efficacy against infection (*VE*_*in*_) = 70%, vaccine efficacy against progression to symptoms (*VE*_*pr*_) = 0%, B) *VE*_*in*_ = 50%, *VE*_*pr*_ = 40%, C) *VE*_*in*_ = 0%, *VE*_*pr*_ = 70%. IRR = incidence rate ratio.

Imperfect test sensitivity and specificity biased efficacy estimates towards zero. Factors increasing the magnitude of the bias were: reduced specificity, reduced sensitivity, increased testing frequency, and calculation with the CIR instead of the IRR. Although the serology estimated *VE*_*in*_ was based on the CIR (as person-time at risk is unknown), the bias was usually lower than the weekly-PCR estimate, for a given sensitivity and specificity, due to the lower frequency of testing (Figure 3).

**Figure 3.**
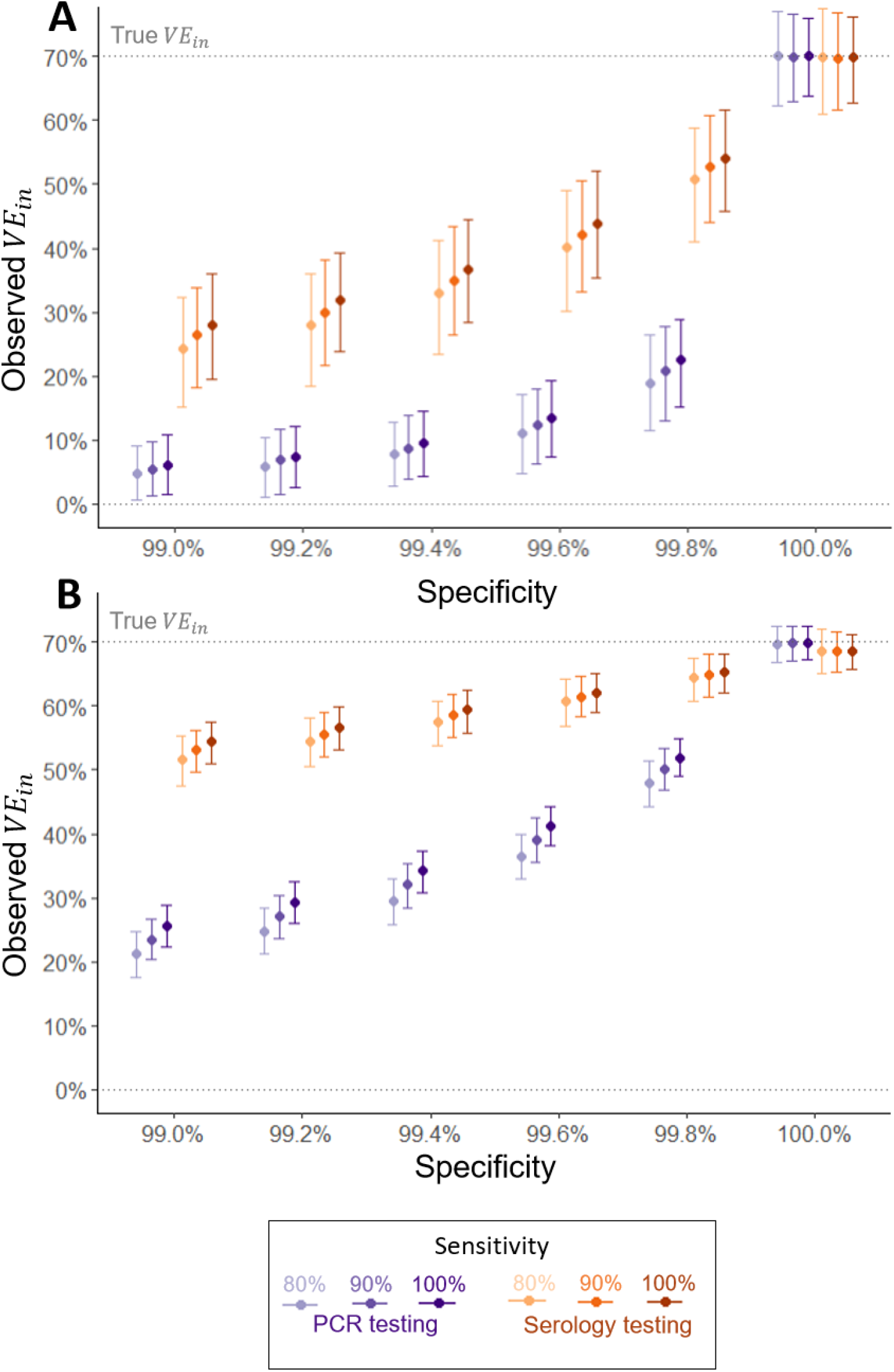
Impact of imperfect test sensitivity and specificity on serology-estimated and PCR-estimated vaccine efficacy against infection. A) Low force of infection (5% per year), B) High force of infection (30% per year). 6-month follow-up visit: Serology tests taken at month 1, 2 and 6 (cumulative seroconversions up to 6-month visit); PCR tests taken weekly. Serology efficacy calculated using 1-CIR; PCR efficacy calculated using 1-IRR. Sensitivity assumed to be equal for symptomatic and asymptomatic infections. Points and error bars represent the mean and 2.5 and 97.5 percentiles from 1000 simulations. *VE*_*in*_= vaccine efficacy against infection, CIR = cumulative incidence ratio, IRR = incidence rate ratio.

For a vaccine that was protective against symptom development, *VE*_*in*_ was overestimated when asymptomatic infections were less likely to be detected than symptomatic infections (Figure 4). The greater the difference in the probability of detection and the greater the vaccine’s protection against symptoms, the greater the overestimation.

**Figure 4.**
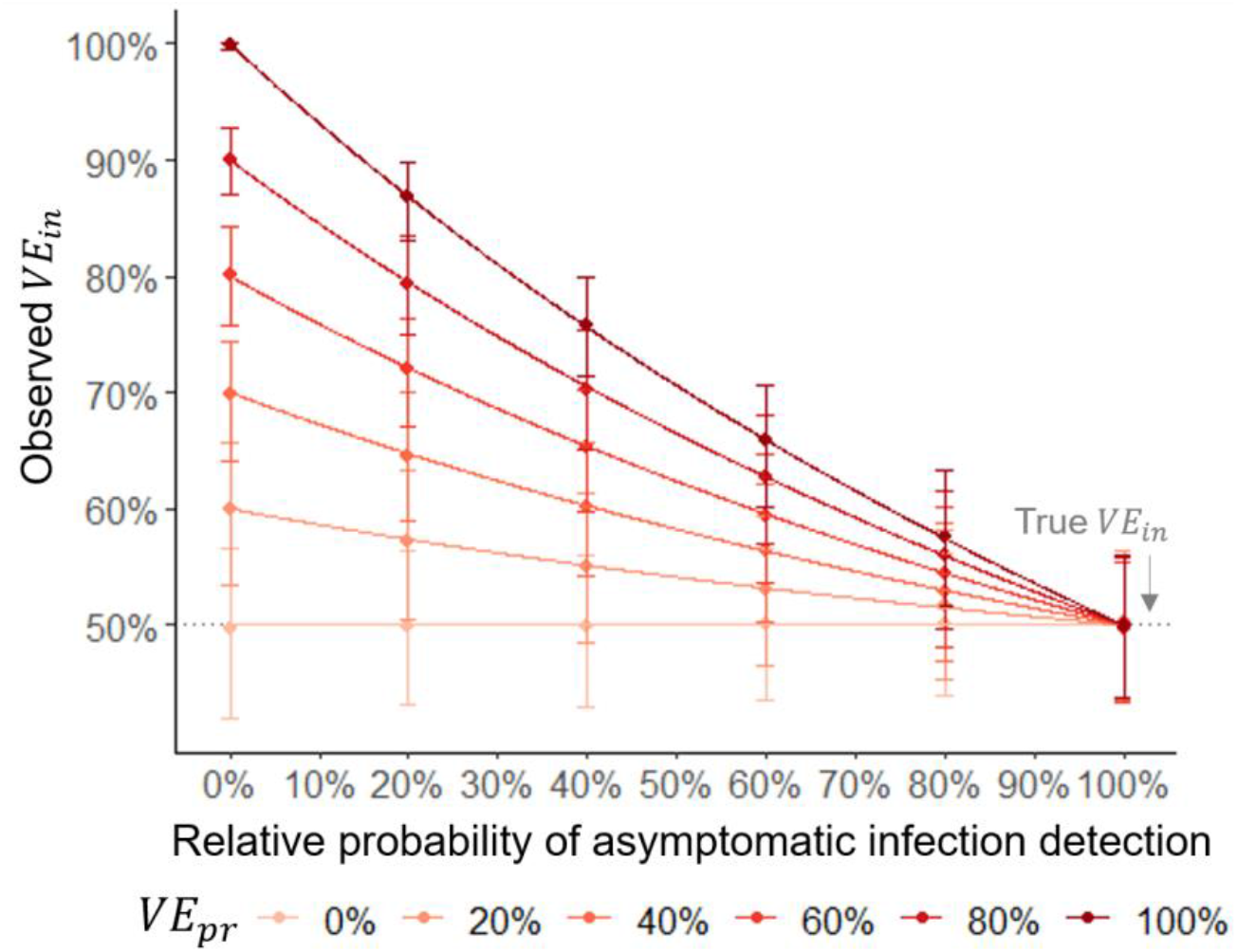
Impact of differential detection of asymptomatic and symptomatic infections on observed vaccine efficacy against infection. Lines show values estimated with Equation 4, points and error bars show the observed mean and 2.5 and 97.5 percentiles from 1000 simulations, with efficacy calculated as 1-IRR, censoring after the first infection. Specificity = 100%, yearly force of infection = 5%, follow up = 12 months from 2 weeks post 2^nd^ dose. *VE*_*in*_= vaccine efficacy against infection, *VE*_*pr*_= vaccine efficacy against progression to symptoms, IRR = incidence rate ratio.

These results were insensitive to adding age stratification to the probability of symptoms. However, also adding age-stratification to vaccine efficacy led to biased estimates of *VE*_*sym*_ and *VE*_*asym*_, when not adjusted for age (e.g. in Poisson regression; Figure 5). When vaccine efficacy decreased with age and the probability of symptoms increased, *VE*_*asym*_ was overestimated and *VE*_*sym*_ underestimated. The magnitude of the difference was greater with an increased association between age and the probability of symptoms, and between age and efficacy.

**Figure 5.**
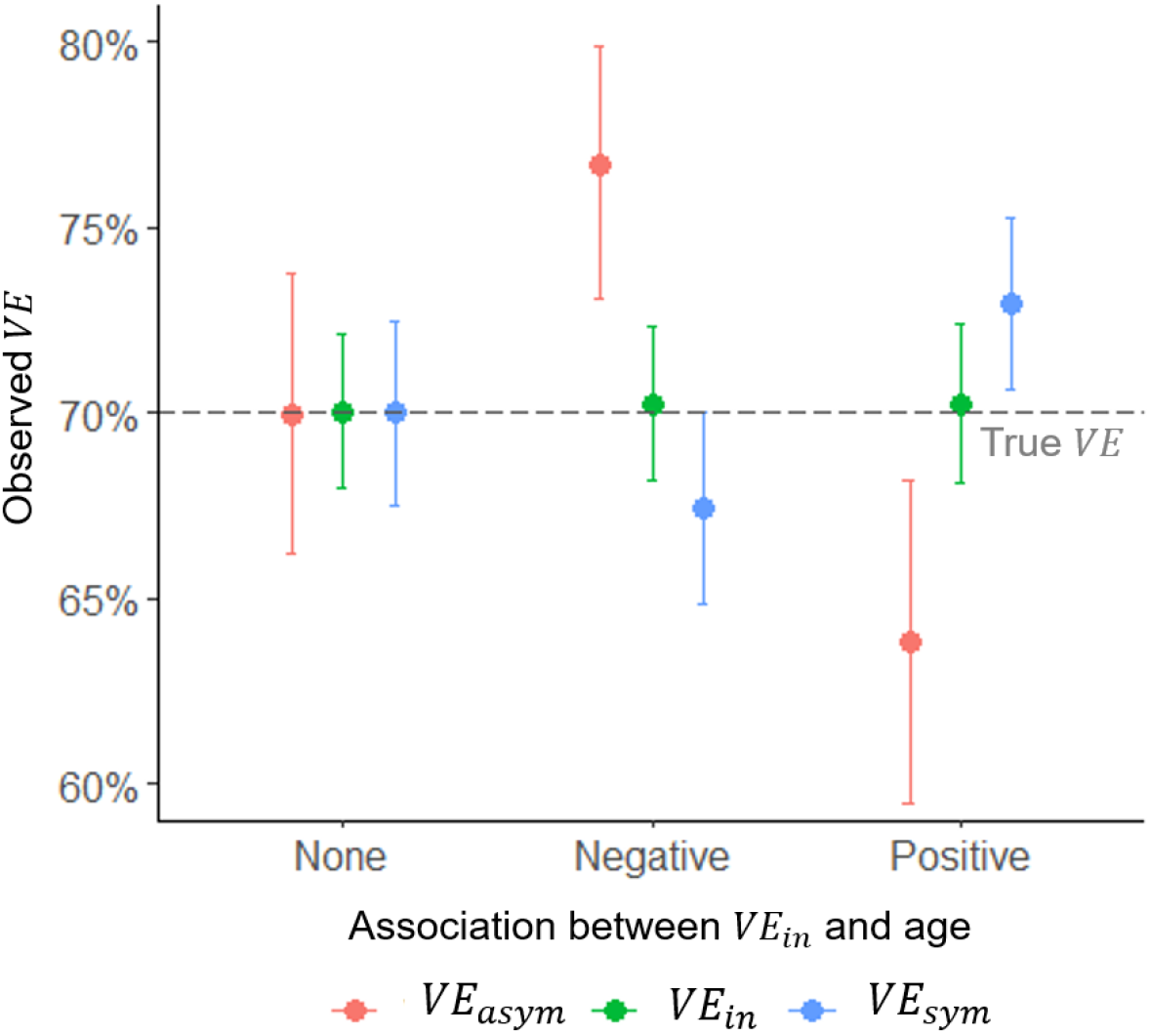
Estimated vaccine efficacy in a population with a higher probability of symptoms with age. Points and error bars represent the mean and 2.5 and 97.5 percentiles of 1000 simulations. True vaccine efficacy against asymptomatic infection (*VE*_*asym*_) = true vaccine efficacy against infection (*VE*_*in*_) = true vaccine efficacy against symptomatic infection (*VE*_*sym*_) = 70%. Specificity = 100%, yearly force of infection = 20%, follow up = 12 months from 2 weeks post 2^nd^ dose.

### Estimating *VE*_*in*_, *VE*_*pr*,_ and the likely bias from the published trial results

Applying Equations 3 and 4 to the reported trial results gave an estimated *VE*_*pr*_ for ChAdOx1 of 43.6% (95% CI 20.6 to 59.9) (Table 2). For Ad26.COV2.S, *VE*_*in*_ was estimated at 66.2% (95% CI 55.9 to 74.1) and *VE*_*pr*_ just 0.9% (95% CI -46.8 to 33.2).

**Table 2.**
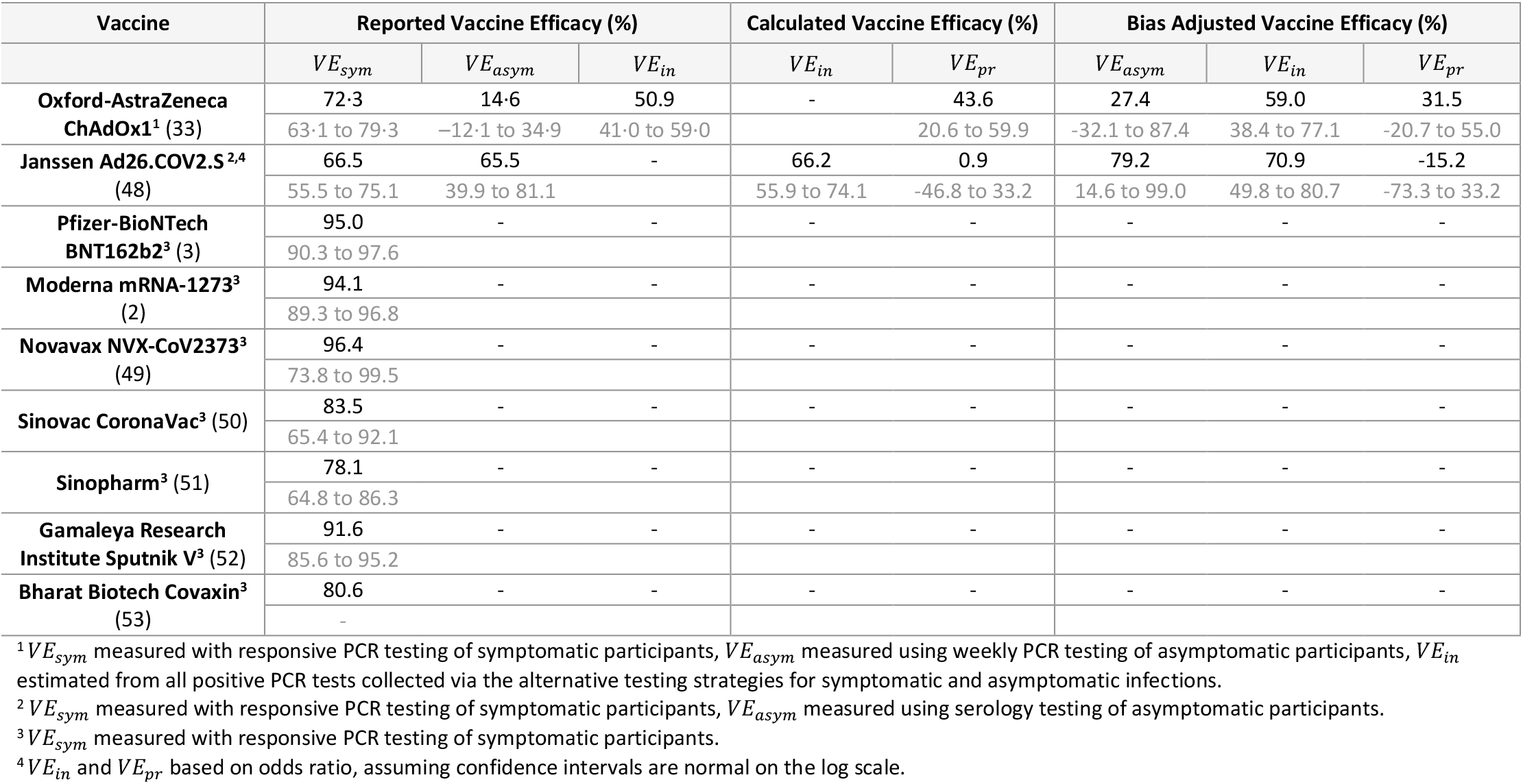
**Estimated vaccine efficacy against infection (*VE***_***in***_**) and vaccine efficacy against progression to symptoms (*VE***_***pr***_**) using trial-reported vaccine efficacy against symptomatic infection (*VE***_***sym***_**) and vaccine efficacy against asymptomatic infection (*VE***_***asym***_). Trials that have not yet reported an estimate are left blank. *VE*_*in*_ and *VE*_*pr*_ calculated using Equations 3 and 4, respectively.

Incorporating the aforementioned biases, the model gave best estimates for ChAdOx1 *VE*_*in*_, *VE*_*asym*_ and *VE*_*pr*_ of 59.0% (95% UI 38.4 to 77.1), 27.4% (95% UI -32.1 to 87.4) and 31.5% (95% UI -20.7 to 55.0) respectively. While for Ad26.COV2.S, the corresponding bias-adjusted estimates were 70.9% (95% CI 49.8 to 80.7), 79.2% (95% CI 14.6 to 99.0) and -15.2% (95% CI -73.3 to 33.2). The rank regression analyses showed that the biases were most sensitive to the test specificity, baseline hazard of infection and adherence to weekly PCR testing (Supplementary Tables 3-5).

## Discussion

Accurately estimating COVID-19 vaccine efficacy against infection and disease is important to understand vaccine benefits, their likely impact on transmission and the long-term prospects for disease control. Estimating the magnitude and direction of biases in a simulated COVID-19 vaccine trial helps to understand their likely influence, and may help to explain differences seen between vaccines, trials, and populations.

We firstly derived the relationship between *VE*_*asym*_ with efficacy against infection and against disease in breakthrough infections. While increasing *VE*_*in*_ increased *VE*_*asym*_, increasing *VE*_*pr*_ had the opposite effect because more infections were prevented from becoming symptomatic. This influence of *VE*_*pr*_ was stronger when a greater proportion of infections were symptomatic in the absence of vaccination. Therefore, although counterintuitive, for COVID-19 where a minority of infections present asymptomatically and the vaccines have high efficacies against symptomatic infection, protection against infection can be expected even when *VE*_*asym*_ is low or negative.

Secondly, we estimated that the ChAdOx1 weekly PCR-measured *VE*_*in*_ was underestimated by 8.1% (Trial 50.9%, Model 59.0%) and *VE*_*asym*_ by 12.8% (Trial 14.6%, Model 27.4%). The *VE*_*pr*_ calculated from the trial reported *VE*_*in*_ and *VE*_*sym*_ would therefore be an overestimation (Calculation 43.6%, Model 31.5%). However, a wide range of values are compatible with the reported trial results when considering stochastic variation and parameter uncertainty. The true *VE*_*in*_ may range between 38.4% and 77.1%, and *VE*_*pr*_ between -20.7% and 55.0%. Given the strong bias towards zero that can be caused by reduced test specificity and a high frequency of testing, it would not be unreasonable for the true *VE*_*in*_ to be closer to our upper uncertainty interval. For Ad26.COV2.S, our model suggests that the true *VE*_*in*_ lies between 49.8% and 80.7%, with a best estimate of 70.9%. Although this indicates a negative *VE*_*pr*_, we believe this is unlikely and rather explained by the small sample size informing the trial reported *VE*_*asym*_ estimate.

We explain these overall expected differences by four biases that are likely to act in the COVID-19 trials.

1. **A lower probability of detecting asymptomatic infections relative to symptomatic infections leads to overestimation of *VE***_***in***_ **if the vaccine protects against progression to symptoms**. For these vaccines (with *VE*_*pr*_ > 0) some infections will be prevented from causing symptoms, so will be less likely to be detected. *VE*_*pr*_ would be mistaken for *VE*_*in*_, so *VE*_*in*_ would be overestimated. Both conditions for this bias are likely to be satisfied in the COVID-19 trials, as multiple studies have indicated that virological and serological testing approaches are less sensitive to asymptomatic infections (20,21). This bias is likely to have influenced the ChAdOx1 *VE*_*in*_ estimate, however we expect it was overridden by a competing downwards bias.
2. **Imperfect test sensitivity and specificity bias estimates towards zero, with greater bias with higher frequency of testing, lower force of infection and when vaccine efficacy is calculated using cumulative incidence rather than incidence rates**. This is caused by a build-up of false positives in both trial arms, and is greater when the ratio of false positives to true positives is greater (8). Regression analysis showed that this was the predominant factor leading to underestimation for both ChAdOx1 and Ad26.COV2.S in our model. As the bias is greater when testing is frequent, even a test with high specificity could bias the estimated ChAdOx1 *VE*_*in*_ and *VE*_*asym*_ noticeably towards zero. This could therefore explain such contrasting trial reported *VE*_*asym*_ estimates between ChAdOx1 and Ad26.COV2.S, despite their similar platforms and neutralising antibody responses (22,23).
3. **A build-up of natural immunity from undetected asymptomatic infections contributes a small downwards bias in *VE***_***sym***_ **for infection-blocking vaccines and a small upwards bias for disease-blocking vaccines**. For an infection-blocking vaccine, the proportion of infections that are asymptomatic is unaltered by the vaccine. Therefore, the rate at which immunity from asymptomatic infections accumulates is equivalent across trial arms, leading to an underestimation of *VE*_*sym*_ (10,24– 26). Yet for disease-blocking vaccines, a greater proportion of infections in the vaccine arm will be asymptomatic, accelerating the acquisition of immunity from undetected infections and introducing a conflicting upward bias. Our model and real-world effectiveness studies suggest the COVID-19 vaccines protect against both infection and symptoms, to varying degrees (27,28). Therefore we expect the overall direction of this bias to be towards zero, and for its magnitude to be greater for vaccines with higher *VE*_*in*_ (e.g., Pfizer-BioNTech BNT162b2 (29)).
4. **Decreasing vaccine efficacy by age will bias estimated *VE***_***sym***_ **downwards and *VE***_***asym***_ **upwards, unless adjusted for age**. This is due to older participants contributing more to *VE*_*sym*_ estimates than younger participants, who contribute more to *VE*_*asym*_ estimates. This bias is dependent on the probability of symptoms increasing with age, for which there is mixed evidence (30–32). However, it should be considered when interpreting estimates based on different subgroups, such as if *VE*_*asym*_ is estimated from a subgroup with serological data while *VE*_*sym*_ is based on the total population.

These biases also apply to effectiveness studies, based on cohort or case-control designs. Most notably, the bias arising from differential detection of asymptomatic and symptomatic infections will likely be greater in real-world studies, where asymptomatic testing is less rigorous. This should be considered when comparing real-world and trial reported estimates, as it could lead to greater bias towards overestimation of *VE*_*in*_ in effectiveness studies, if the vaccine prevents symptom development.

There are limitations to our analysis, notably uncertainties over parameter estimates. There is limited evidence on both serology and PCR test sensitivities for asymptomatic infections, and how these change over time since infection. As we show, differences in test sensitivity by symptom status can lead to overestimation of *VE*_*in*_, so further studies are needed to clarify the potential role of this bias. We also did not consider the vaccines’ effects on viral load and how this alters virological and serological test sensitivity. Multiple COVID-19 vaccines have been shown to reduce SARS-CoV-2 viral load (33,34), and lower load infections are less likely to lead to seroconversion (35). Therefore serology-based efficacy estimates may be more representative of high viral load infections than all infections. They may be comparable to estimates based on DNA sequenced swabs, as these samples must exceed a threshold viral load to be sequenced. Finally, we do not consider the use of point prevalence estimates from single time point PCR swabs, however this has been explored elsewhere (11,12).

In conclusion, multiple biases have the potential to influence the COVID-19 vaccine efficacy estimates, with their direction and magnitude dependent on the vaccine properties and testing strategies. These biases may explain differences between the ChAdOx1 and Ad26.COV2.S trial reported estimates despite similar vaccine platform technologies, and should be considered when interpreting both efficacy and effectiveness study results as these begin to be reported for these and other vaccines.

## Data Availability

Code is available on GitHub.

https://github.com/lucyrose96/COVID-19-Trial-Model

## References

1. Voysey M, Costa Clemens SA, Madhi SA, Weckx LY, Folegatti PM, Aley PK, et al. Single-dose administration and the influence of the timing of the booster dose on immunogenicity and efficacy of ChAdOx1 nCoV-19 (AZD1222) vaccine: a pooled analysis of four randomised trials. Lancet. 2021;397(10277):881–91.

2. Baden LR, El Sahly HM, Essink B, Kotloff K, Frey S, Novak R, et al. Efficacy and Safety of the mRNA-1273 SARS-CoV-2 Vaccine. N Engl J Med. 2021;384(5):403–16.

3. Polack FP, Thomas SJ, Kitchin N, Absalon J, Gurtman A, Lockhart S, et al. Safety and Efficacy of the BNT162b2 mRNA Covid-19 Vaccine. N Engl J Med. 2020;383(27):2603–15.

4. Hodgson SH, Mansatta K, Mallett G, Harris V, Emary KRW, Pollard AJ. What defines an efficacious COVID-19 vaccine? A review of the challenges assessing the clinical efficacy of vaccines against SARS-CoV-2. Vol. 21, The Lancet Infectious Diseases. 2021. p. e26–35.

5. Moore S, Hill EM, Tildesley MJ, Dyson L, Keeling MJ. Vaccination and non-pharmaceutical interventions for COVID-19: a mathematical modelling study. Lancet Infect Dis. Elsevier BV; 2021 Mar;21(6):793–802.

6. Rapaka RR, Hammershaimb EA, Neuzil KM. Are some COVID vaccines better than others? Interpreting and comparing estimates of efficacy in trials of COVID-19 vaccines. Clin Infect Dis. 2021;

7. Lipsitch M, Dean NE. Understanding COVID-19 vaccine efficacy. Vol. 370, Science. 2020. p. 763–5.

8. Lachenbruch PA. Sensitivity, specificity, and vaccine efficacy. Control Clin Trials. 1998;19(6):569–74.

9. Kahn R, Hitchings M, Wang R, Bellan SE, Lipsitch M. Analyzing Vaccine Trials in Epidemics with Mild and Asymptomatic Infection. Am J Epidemiol. 2019;188(2):467–74.

10. Smith PG, Rodrigues LC, Fine PEM. Assessment of the protective efficacy of vaccines against common diseases using case-control and cohort studies. Int J Epidemiol. 1984;13(1):87–93.

11. Lipsitch M, Kahn R. Interpreting vaccine efficacy trial results for infection and transmission. Vaccine. Cold Spring Harbor Laboratory Preprints; 2021 Feb 28;39(30):4082–8.

12. Follmann D, Fay M. Vaccine Efficacy at a Point in Time. medRxiv. 2021 Jan 1;2021.02.04.21251133.

13. Halloran ME, Longini IM, Struchiner CJ. Introduction and Examples. In: Design and Analysis of Vaccine Studies. Springer-Verlag New York; 2010. p. 1–18.

14. Halloran ME, Longini IM, Struchiner CJ. Overview of Vaccine Effects and Study Designs. In: Design and Analysis of Vaccine Studies. Springer-Verlag New York; 2010. p. 19–45.

15. Oran DP, Topol EJ. The Proportion of SARS-CoV-2 Infections That Are Asymptomatic:A Systematic Review. Ann Intern Med. American College of Physicians; 2021 Jan 22;174(5):655– 62.

16. Ainsworth M, Andersson M, Auckland K, Baillie JK, Barnes E, Beer S, et al. Performance characteristics of five immunoassays for SARS-CoV-2: a head-to-head benchmark comparison. Lancet Infect Dis. 2020;20(12):1390–400.

17. Harritshøj LH, Gybel-Brask M, Afzal S, Kamstrup PR, Jørgensen CS, Thomsen MK, et al. Comparison of 16 serological SARS-CoV-2 immunoassays in 16 clinical laboratories. J Clin Microbiol. 2021;59(5):2596–616.

18. Hellewell J, Russell TW, Beale R, Kelly G, Houlihan C, Nastouli E, et al. Estimating the effectiveness of routine asymptomatic PCR testing at different frequencies for the detection of SARS-CoV-2 infections. BMC Med. BioMed Central; 2021 Apr 27;19(1):106.

19. Skittrall JP, Wilson M, Smielewska AA, Parmar S, Fortune MD, Sparkes D, et al. Specificity and positive predictive value of SARS-CoV-2 nucleic acid amplification testing in a low-prevalence setting. Clin Microbiol Infect. Elsevier B.V.; 2021;27(3):469.e9-469.e15.

20. Lumley SF, Wei J, O’Donnell D, Stoesser NE, Matthews PC, Howarth A, et al. The duration, dynamics and determinants of SARS-CoV-2 antibody responses in individual healthcare workers. Clin Infect Dis. 2021;

21. Long QX, Tang XJ, Shi QL, Li Q, Deng HJ, Yuan J, et al. Clinical and immunological assessment of asymptomatic SARS-CoV-2 infections. Nat Med. 2020;26(8):1200–4.

22. Folegatti PM, Ewer KJ, Aley PK, Angus B, Becker S, Belij-Rammerstorfer S, et al. Safety and immunogenicity of the ChAdOx1 nCoV-19 vaccine against SARS-CoV-2: a preliminary report of a phase 1/2, single-blind, randomised controlled trial. Lancet. 2020;396(10249):467–78.

23. Sadoff J, Le Gars M, Shukarev G, Heerwegh D, Truyers C, de Groot AM, et al. Interim Results of a Phase 1–2a Trial of Ad26.COV2.S Covid-19 Vaccine. N Engl J Med. 2021;384(19):1824–35.

24. Wu Y, Marsh JA, McBryde ES, Snelling TL. The influence of incomplete case ascertainment on measures of vaccine efficacy. Vaccine. 2018;36(21):2946–52.

25. Lipsitch M. Challenges of vaccine effectiveness and waning studies. Vol. 68, Clinical Infectious Diseases. 2019. p. 1631–3.

26. Lewnard JA, Tedijanto C, Cowling BJ, Lipsitch M. Measurement of vaccine direct effects under the test-negative design. Am J Epidemiol. 2018;187(12):2686–97.

27. Menni C, Klaser K, May A, Polidori L, Capdevila J, Louca P, et al. Vaccine side-effects and SARS-CoV-2 infection after vaccination in users of the COVID Symptom Study app in the UK: a prospective observational study. Lancet Infect Dis. 2021;21(7):939–49.

28. Haas EJ, Angulo FJ, McLaughlin JM, Anis E, Singer SR, Khan F, et al. Impact and effectiveness of mRNA BNT162b2 vaccine against SARS-CoV-2 infections and COVID-19 cases, hospitalisations, and deaths following a nationwide vaccination campaign in Israel: an observational study using national surveillance data. Lancet. Lancet; 2021 May 5;397(10287):1819–29.

29. Thompson MG, Burgess JL, Naleway AL, Tyner HL, Yoon SK, Meece J, et al. Interim Estimates of Vaccine Effectiveness of BNT162b2 and mRNA-1273 COVID-19 Vaccines in Preventing SARS-CoV-2 Infection Among Health Care Personnel, First Responders, and Other Essential and Frontline Workers — Eight U.S. Locations, December 2020–March. MMWR Morb Mortal Wkly Rep. 2021;70(13):495–500.

30. Poletti P, Tirani M, Cereda D, Trentini F, Guzzetta G, Sabatino G, et al. Association of Age with Likelihood of Developing Symptoms and Critical Disease among Close Contacts Exposed to Patients with Confirmed SARS-CoV-2 Infection in Italy. JAMA Netw Open. American Medical Association; 2021 Mar 10;4(3):211085.

31. Davies NG, Klepac P, Liu Y, Prem K, Jit M, Pearson CAB, et al. Age-dependent effects in the transmission and control of COVID-19 epidemics. Nat Med. Nature Research; 2020 Aug 1;26(8):1205–11.

32. Elliott J, Whitaker M, Bodinier B, Riley S, Ward H, Cooke G, et al. Symptom reporting in over 1 million people: community detection of COVID-19. medRxiv. 2021;2021.02.10.21251480.

33. Emary KRW, Golubchik T, Aley PK, Ariani C V., Angus B, Bibi S, et al. Efficacy of ChAdOx1 nCoV-19 (AZD1222) vaccine against SARS-CoV-2 variant of concern 202012/01 (B.1.1.7): an exploratory analysis of a randomised controlled trial. Lancet. 2021;397(10282):1351–62.

34. Pritchard E, Matthews PC, Stoesser N, Eyre DW, Gethings O, Vihta KD, et al. Impact of vaccination on new SARS-CoV-2 infections in the United Kingdom. Nat Med. 2021;

35. Masiá M, Telenti G, Fernández M, García JA, Agulló V, Padilla S, et al. SARS-CoV-2 Seroconversion and Viral Clearance in Patients Hospitalized with COVID-19: Viral Load Predicts Antibody Response. Open Forum Infect Dis. 2021;8(2).

36. ClinicalTrials.gov [Internet]. Bethesda (MD): National Library of Medicine (US). 2000 Feb 29 -. Identifier NCT04516746, Phase III Double-blind, Placebo-controlled Study of AZD1222 for the Prevention of COVID-19 in Adults; 2020 Aug 18 [cited 2021 Jul 27]. Available from: https://clinicaltrials.gov/ct2/show/NCT04516746

37. ClinicalTrials.gov [Internet]. Bethesda (MD): National Library of Medicine (US). 2000 Feb 29 -. Identifier NCT04505722, A Study of Ad26.COV2.S for the Prevention of SARS-CoV-2-Mediated COVID-19 in Adult Participants (ENSEMBLE); 2020 Aug 10 [cited 2021 Jul 27]. Available from: https://clinicaltrials.gov/ct2/show/study/NCT04505722

38. ClinicalTrials.gov [Internet]. Bethesda (MD): National Library of Medicine (US). 2000 Feb 29 -. Identifier NCT04614948, A Study of Ad26.COV2.S for the Prevention of SARS-CoV-2-mediated COVID-19 in Adults (ENSEMBLE 2); 2020 Nov 4 [cited 2021 Jul 27]. Available from: https://clinicaltrials.gov/ct2/show/NCT04614948

39. ClinicalTrials.gov [Internet]. Bethesda (MD): National Library of Medicine (US). 2000 Feb 29 -. Identifier NCT04530396, Clinical Trial of Efficacy, Safety, and Immunogenicity of Gam-COVID-Vac Vaccine Against COVID-19 (RESIST); 2020 Aug 28 [cited 2021 Jul 27]. Available from: https://clinicaltrials.gov/ct2/show/NCT04530396

40. ClinicalTrials.gov [Internet]. Bethesda (MD): National Library of Medicine (US). 2000 Feb 29 -. Identifier NCT04368728, Study to Describe the Safety, Tolerability, Immunogenicity, and Efficacy of RNA Vaccine Candidates Against COVID-19 in Healthy Individuals; 2020 Apr 30 [cited 2021 Jul 27]. Available from: https://clinicaltrials.gov/ct2/show/NCT04368728?term=AREA%5BInterventionType%5D+%28Drug+OR+Biological%29&recrs=a&cond=COVID-19&intr=vaccine&phase=2&draw=2

41. ClinicalTrials.gov [Internet]. Bethesda (MD): National Library of Medicine (US). 2000 Feb 29 -. Identifier NCT04470427, A Study to Evaluate Efficacy, Safety, and Immunogenicity of mRNA-1273 Vaccine in Adults Aged 18 Years and Older to Prevent COVID-19; 2020 Jul 14 [cited 2021 Jul 27]. Available from: https://clinicaltrials.gov/ct2/show/NCT04470427

42. ClinicalTrials.gov [Internet]. Bethesda (MD): National Library of Medicine (US). 2000 Feb 29 -. Identifier NCT04583995, A Study Looking at the Effectiveness, Immune Response, and Safety of a COVID-19 Vaccine in Adults in the United Kingdom; 2020 Oct 12 [cited 2021 Jul 27]. Available from: https://clinicaltrials.gov/ct2/show/NCT04583995

43. ClinicalTrials.gov [Internet]. Bethesda (MD): National Library of Medicine (US). 2000 Feb 29 -. Identifier NCT04611802, A Study to Evaluate the Efficacy, Immune Response, and Safety of a COVID-19 Vaccine in Adults ≥ 18 Years With a Pediatric Expansion in Adolescents (12 to < 18 Years) at Risk for SARS-CoV-2; 2020 Nov 2 [cited 2021 Jul 27]. Available from: https://clinicaltrials.gov/ct2/show/NCT04611802

44. ClinicalTrials.gov [Internet]. Bethesda (MD): National Library of Medicine (US). 2000 Feb 29 -. Identifier NCT04641481, An Efficacy and Safety Clinical Trial of an Investigational COVID-19 Vaccine (BBV152) in Adult Volunteers; 2020 Nov 23 [cited 2021 Jul 27]. Available from: https://clinicaltrials.gov/ct2/show/NCT04641481

45. ClinicalTrials.gov [Internet]. Bethesda (MD): National Library of Medicine (US). 2000 Feb 29 -. Identifier NCT04510207, A Study to Evaluate The Efficacy, Safety and Immunogenicity of Inactivated SARS-CoV-2 Vaccines (Vero Cell) in Healthy Population Aged 18 Years Old and Above (COVID-19); 2020 Aug 12 [cited 2021 Jul 27]. Available from: https://clinicaltrials.gov/ct2/show/NCT04510207

46. ClinicalTrials.gov [Internet]. Bethesda (MD): National Library of Medicine (US). 2000 Feb 29 -. Identifier NCT04456595, Clinical Trial of Efficacy and Safety of Sinovac’s Adsorbed COVID-19 (Inactivated) Vaccine in Healthcare Professionals (PROFISCOV); 2020 Jul 2 [cited 2021 Jul 27]. Available from: https://clinicaltrials.gov/ct2/show/NCT04456595

47. ClinicalTrials.gov [Internet]. Bethesda (MD): National Library of Medicine (US). 2000 Feb 29 -. Identifier NCT04400838, Investigating a Vaccine Against COVID-19; 2020 May 26 [cited 2021 Jul 27]. Available from: https://clinicaltrials.gov/ct2/show/NCT04400838

48. Sadoff J, Gray G, Vandebosch A, Cárdenas V, Shukarev G, Grinsztejn B, et al. Safety and Efficacy of Single-Dose Ad26.COV2.S Vaccine against Covid-19. N Engl J Med. Massachusetts Medical Society; 2021 Apr 21;384(23):2187–201.

49. Heath PT, Galiza EP, Baxter DN, Boffito M, Browne D, Burns F, et al. Safety and Efficacy of NVX-CoV2373 Covid-19 Vaccine. N Engl J Med. Massachusetts Medical Society; 2021 Jun 30;NEJMoa2107659.

50. Tanriover MD, Doğanay HL, Akova M, Güner HR, Azap A, Akhan S, et al. Efficacy and safety of an inactivated whole-virion SARS-CoV-2 vaccine (CoronaVac): interim results of a double-blind, randomised, placebo-controlled, phase 3 trial in Turkey. Lancet. Elsevier; 2021 Jul 17;398(10296):213–22.

51. Al Kaabi N, Zhang Y, Xia S, Yang Y, Al Qahtani MM, Abdulrazzaq N, et al. Effect of 2 Inactivated SARS-CoV-2 Vaccines on Symptomatic COVID-19 Infection in Adults: A Randomized Clinical Trial. JAMA - J Am Med Assoc. American Medical Association; 2021 Jul 6;326(1):35–45.

52. Logunov DY, Dolzhikova I V., Shcheblyakov D V., Tukhvatulin AI, Zubkova O V., Dzharullaeva AS, et al. Safety and efficacy of an rAd26 and rAd5 vector-based heterologous prime-boost COVID-19 vaccine: an interim analysis of a randomised controlled phase 3 trial in Russia. Lancet. Elsevier B.V.; 2021 Feb 20;397(10275):671–81.

53. Bharat Biotech. Bharat Biotech Announces Phase 3 Results of COVAXIN®: India’s First COVID-19 Vaccine Demonstrates Interim Clinical Efficacy of 81% [Internet]. 2021 [cited 2021 Jul 26]. Available from: https://www.bharatbiotech.com/images/press/covaxin-phase3-efficacy-results.pdf

