## Supplementary Materials for "Measuring vaccine efficacy against infection and disease in clinical trials: sources and magnitude of bias in COVID-19 vaccine efficacy estimates"

### Supplementary Methods

#### ***Mathematical Model for COVID-19 vaccine trials***

We assume that the duration of infection is described by an Erlang distribution (rate = 0.724 per day, shape = 12), giving a mean of 16 days and standard deviation of 4.8 days (Supplementary Figure 3B). Upon recovery, trial participants are assumed to produce detectable antibodies (seroconvert) and to develop natural immunity. This natural immunity is assumed to provide 84% protection from infection (1) and 65% additional protection from developing symptoms (2). We assume that once a vaccinated participant has experienced a natural infection, the vaccine provides no additional protection.

Time to loss of detectable antibodies (seroreversion) is described by an exponential distribution with a mean of 110 months (Supplementary Figure 3C) (3). Due to the limited evidence on protective immunity following seroreversion after natural infection, we assume continued protection against infection at the same level, but perform sensitivity analysis to this assumption. Seropositive participants who are reinfected stay seropositive throughout their infection, while participants who have seroreverted seroconvert following reinfection at a faster rate than naïve individuals, to reflect immune memory. This is given by an Erlang distribution (rate = 0.724 per day, shape = 5), giving a mean of 4.3 days to seroreversion for reinfections in seronegative participants (Supplementary Figure 3D).

We define the start of follow-up as two weeks post final dose. In line with the published trial protocols, serological testing is performed at 1, 2, 6, 12 and 24 months after the baseline. Seropositive participants whose most recent infection was symptomatic are detected with a probability of 95%, while asymptomatic infections are detected with a probability of 80% (4). At each test, seronegative participants are detected with a probability of 1-specificity, where specificity is assumed to be 99.84% (5).

We assume PCR-testing of all trial participants every 7 days, as performed in the Oxford-AstraZeneca UK trial (COV002) (6). The specificity of PCR-testing is estimated at 99.945% (7), and the per test probability of a false positive is given by 1-specificity. While we acknowledge that some false positives will occur in participants that have recently reported symptoms consistent with a COVID-19 diagnosis, we expect this to be negligible.

To align with the published estimates (8,9), we ran the model for 149 days for Oxford-AstraZeneca ChAdOx1 and estimated the observed $VE_{asym}$ using the incidence of asymptomatic infections detected with weekly PCR testing, the observed $VE_{sym}$ using the incidence of symptomatic infections, and the observed $VE_{in}$ from the sum of the detected asymptomatic and symptomatic infections. We estimated the true $VE_{in}$ and $VE_{asym}$ as the mean parameter values that gave observed values corresponding with the trial reported estimates. We calculated the true $VE_{pr}$ from the bias-adjusted $VE_{in}$ and $VE_{sym}$. For Janssen Ad26.COV2.S, we ran the model for 71 days and calculated $VE_{asym}$ from the incidence of asymptomatic infections detected with serology after day 29. We estimated the true $VE_{asym}$ as the mean parameter value that gave observed values corresponding with the trial reported estimate, then calculated the true $VE_{pr}$ from the bias-adjusted $VE_{asym}$ and $VE_{sym}$.

All analyses were performed in R version 4.0.3.

***Estimating the number of symptomatic false positives***

We estimate the expected number of symptomatic false positives using the following formula

$$N_{se} \times (1 - Sp^{N_t})$$

where $N_{se}$ represents the number episodes of COVID-19 symptoms without SARS-CoV-2 infection, $Sp$ represents the test specificity and $N_t$, the number of tests around the period of symptoms.

We apply this to the Pfizer estimates for their safety population at the time of the FDA approval application, where 2714 of 37586 (7.2%) participants experienced symptoms that were consistent with COVID-19 but were not confirmed cases, from one week post-dose to a median of two months ("suspected COVID-19 cases") (10). If we assume that i) each of these symptomatic cases had one symptomatic episode during this time, ii) on average, four tests are taken around the symptomatic period (likely to be a maximum), and iii) PCR-test specificity is 99.98% (7) we can estimate that approximately $2714 \times (1 - 0.9998^4) = 2.2$ symptomatic false positives would occur over 2 months follow up in a population of 37586. Extrapolating to 24 months, we estimate approximately $24 \times \frac{2714}{2} \times (1 - 0.9998^4) = 26$ false positives would occur in SARS-CoV-2 negative participants who have symptoms that are consistent with COVID-19, over 24 months of follow up, in a population of 37586. We ignore the censoring of true positives, and the possibility for multiple symptomatic false positives, which would both decrease the number of symptomatic false positive cases. We therefore make a simplifying assumption in our model that all false positives are asymptomatic.

**Sensitivity to heterogeneity in probability of symptoms and vaccine efficacy by age**

The model assumes no heterogeneity in population characteristics. However, we performed a sensitivity analysis to assess the effect of variation in $p_s$ and vaccine efficacy by age. We compared the observed vaccine efficacies ($VE_{sym}$, $VE_{asym}$, and $VE_{in}$) for a population with one age group where $p_s$ = 0.67 and $VE_{in}$= 0.7, with three populations with five age groups of equal proportions, with heterogeneity in $p_s$ and/or $VE_{in}$. In all heterogenous populations, $p_s$ increased in units of 0.1 from age group 1 (youngest), where $p_s = 0.1$, to age group 5 (oldest), where $p_s = 0.5$. In one heterogenous population, $VE_{in}$ did not change with age. In another, $VE_{in}$ increased with age in units of 10%, from 50% to 90%. In the last, $VE_{in}$ decreased with age from 90% to 50%. In all populations $VE_{pr}$ = 0.

***Confidence intervals for vaccine efficacy against infection and progression to symptoms, calculated from the trial reported vaccine efficacy against symptomatic and asymptomatic infection.***

To obtain confidence intervals around vaccine efficacy (*VE*), the natural log of the relative risk (*RR*) of the outcome in the vaccine compared with the control arm ($= \ln(1 - VE)$) is typically assumed to be approximately normal. In this case, the 95% confidence intervals on *VE* are given by $1 - \exp[\ln(RR) \pm 1.96\sigma]$, where $\sigma$ is the standard deviation of the $\ln(RR)$ and is typically estimated in a regression model (e.g. Poisson). From the relationship of $VE_{pr}$ with $VE_{in}$ and $VE_{sym}$ defined in the main text, we know that $RR_{pr} = RR_{sym}/RR_{in}$. If we assume that $RR_{sym}$ and $RR_{in}$ are independent, then $\sigma_{pr}^2 = \sigma_{sym}^2 + \sigma_{in}^2$, where $\sigma_{sym}^2$ is the variance of the $\ln(RR)$ of symptomatic disease in the vaccine compared with the control arm, $\sigma_{in}^2$ the same for infection, etc. and confidence intervals can be obtained as above. We derived estimates of these variance terms from reported vaccine efficacies and associated confidence intervals. Where $VE_{in}$ was not reported, we calculated its value using Equation 3. In this case, we derived the variance of $\ln(RR_{in})$ using the delta method and assuming independence of $RR_{sym}$ and $RR_{asym}$,

$$\sigma_{in}^2 = \frac{(1-p_s)^2\sigma_{asym}^2 RR_{asym}^2 + {p_s}^2\sigma_{sym}^2 RR_{sym}^2}{(VE_{in}-1)^2}$$

***Uncertainty intervals for bias-adjusted vaccine efficacy estimates***

To estimate uncertainty intervals that account for uncertainty in model parameters, we performed uncertainty analysis for our estimates of $VE_{in}$, $VE_{asym}$ and $VE_{pr}$ for Oxford-AstraZeneca ChAdOx1and Janssen Ad26.COV2.S. Parameter values were selected from a uniform distribution within the parameter ranges given in Supplementary Table 2.

We assumed that the trial reported $VE_{sym}$ values were accurate, and that serology test sensitivity to symptomatic infections was equal to or greater than that for asymptomatic infections. As a proxy for adherence to weekly asymptomatic PCR testing, we varied the number of days between each asymptomatic PCR test, from 7 (representing 100% adherence) to 14 (representing 50% adherence). The probability of an asymptomatic infection being detected with each frequency of testing was calculated using the same approach as used for weekly testing (Supplementary Table 1), with data from Hellewell *et al.* (2021) (11).

We ran 10,000 simulations and selected those with observed vaccine efficacy estimates within the 95% confidence intervals reported in the trials. The 2.5 and 97.5 percentiles for the true vaccine efficacy values inputted into the model gave the 95% uncertainty intervals for each true vaccine efficacy estimate.

***Rank regression analysis***

To estimate the contribution of factors to biases in vaccine efficacy estimates, we performed rank regression analysis on the 10,000 simulations generated for the estimates for the bias-adjusted uncertainty intervals. The following additive variables were included in both Oxford-AstraZeneca and Janssen regression models: true $VE_{in}$, true $VE_{pr}$, yearly baseline hazard of infection, proportion of infections that are symptomatic, relative risk of infection with prior infection, relative risk of symptoms with prior infection. For Oxford-AstraZeneca, we also added PCR test sensitivity to asymptomatic infections, PCR test specificity, and the number of days between each PCR test. For Janssen, we also added serology test sensitivity to asymptomatic and symptomatic infections, and serology test specificity.

<u>Supplementary Tables</u>

***Supplementary Table 1. Calculation of the probability of asymptomatic infection detection with weekly PCR testing***. Daily probability of detection estimates from Hellewell et al. (2021) (11).

| Days since infection | Probability of detection | First test day 0 | First test day 1 | First test day 2 | First test day 3 | First test day 4 | First test day 5 | First test day 6 | |
|---|---|---|---|---|---|---|---|---|---|
| 0 | 0.49% | ✓ | | | | | | | |
| 1 | 3.95% | | ✓ | | | | | | |
| 2 | 24.60% | | | ✓ | | | | | |
| 3 | 70.99% | | | | ✓ | | | | |
| 4 | 78.54% | | | | | ✓ | | | |
| 5 | 76.67% | | | | | | ✓ | | |
| 6 | 72.51% | | | | | | | ✓ | |
| 7 | 67.75% | ✓ | | | | | | | |
| 8 | 62.60% | | ✓ | | | | | | |
| 9 | 57.12% | | | ✓ | | | | | |
| 10 | 51.47% | | | | ✓ | | | | |
| 11 | 45.87% | | | | | ✓ | | | |
| 12 | 40.40% | | | | | | ✓ | | |
| 13 | 35.09% | | | | | | | ✓ | |
| 14 | 30.19% | ✓ | | | | | | | |
| 15 | 25.70% | | ✓ | | | | | | |
| 16 | 21.66% | | | ✓ | | | | | |
| 17 | 18.02% | | | | ✓ | | | | |
| 18 | 14.90% | | | | | ✓ | | | |
| 19 | 12.24% | | | | | | ✓ | | |
| 20 | 9.99% | | | | | | | ✓ | |
| 21 | 8.16% | ✓ | | | | | | | |
| 22 | 6.65% | | ✓ | | | | | | |
| 23 | 5.38% | | | ✓ | | | | | |
| 24 | 4.35% | | | | ✓ | | | | |
| 25 | 3.51% | | | | | ✓ | | | |
| 26 | 2.81% | | | | | | ✓ | | |
| 27 | 2.26% | | | | | | | ✓ | |
| 28 | 1.81% | ✓ | | | | | | | |
| 29 | 1.45% | | ✓ | | | | | | |
| 30 | 1.16% | | | ✓ | | | | | |
| Probability of detection over course of infection | | 79.80% | 75.44% | 76.32% | 88.96% | 90.46% | 88.14% | 84.30% | **Average 83.35%** |

***Supplementary Table 2. Model parameter estimates.*** Provided for the range of parameter values explored, and the values specified when estimating the direction and magnitude of the bias for the Oxford-AstraZeneca ChAdOx1 in Emary *et al.* 2021 (8), and Janssen Ad26.COV2.S in Sadoff *et al.* 2021 (9).

| | **Scenarios explored**[1] | **Oxford-AstraZeneca ChAdOx1** | | **Janssen Ad26.COV2.S** | |
|---|---|---|---|---|---|
| **Parameter** | Estimate | Estimate (range | Data source | Estimate (range) | Data source |
| Number completing trial | 30,000 | 8534 | (8) | 43,783 | (10) |
| Proportion in vaccine group | 0.50 | 0.50 | (8) | 0.50 | (10) |
| Yearly baseline hazard of infection | 0.05, 0.10, **0.20**, 0.30 | 0.17 (0.10-0.20) | (8) | 0.35 (0.3-0.4) | (10) |
| Probability that an infection is symptomatic (no age stratification) | 0.0, 0.1, 0.2, **0.3**, 0.4, 0.5, 0.6, 0.7, 0.8, 0.9, 1.0 | 0.67 (0.50-0.80) | (12) | 0.67 (0.50-0.80) | (12) |
| Probability that an infection is symptomatic (with age stratification) | 5 age categories: 0.1, 0.2, 0.3, 0.4, 0.5 | - | - | - | - |
| Relative risk of infection after natural infection | 0.16 | 0.16 (0.10-0.20) | (13) | 0.16 (0.10-0.20) | (13) |
| Relative risk of progression to symptoms after natural infection | 0.35[2] | 0.35 (0.30-0.40)[2] | (2) | 0.35 (0.30-0.40)[2] | (2) |
| Vaccine efficacy against infection | 0.0, 0.1, 0.2, 0.3, 0.4, 0.5, 0.6, 0.7, 0.8, 0.9, 1.0 | 0.0 – 0.723[3] | - | 0.0 – 0.665[3] | - |
| Vaccine efficacy against progression to symptoms | 0.0, 0.1, 0.2, 0.3, 0.4, 0.5, 0.6, 0.7, 0.8, 0.9, 1.0 | Min $VE_{pr}$ – 0.723 [3,4] | - | Min $VE_{pr}$ – 0.665 [3,4] | - |
| Number of days between asymptomatic PCR tests[6] | 7 | 7-14 | - | - | - |
| Probability of an asymptomatic infection being detected with weekly PCR testing | 0.0, 0.1, 0.2, 0.3, 0.4, 0.5, 0.6, 0.7, **0.8**, 0.9, 1.0 | 0.8335 (0.650-0.945) | (11) | - | - |
| Adherence to weekly PCR testing | 1.0 | 0.5-1.0 | - | - | - |
| Probability of a symptomatic infection being detected with PCR testing upon reporting of symptoms | 1.00 | 1.00 | - | 1.00 | - |
| PCR specificity | 0.990, 0.992, 0.994, 0.996, 0.998, **1.000** | 0.99945 (0.9991-0.9998) | (7) | 0.99945 (0.99910-0.99980) | (7) |
| Number serology tested | 30,000 | - | - | 2650 | (10) |
| Median time to seroconversion (days from infection) | 16 | - | - | 16 | (14) |
| Serology sensitivity for asymptomatic infections | **0.80**, 0.90, 0.95, 1.00 | - | - | 0.80 (0.6-0.99)[5] | (4) |
| Serology sensitivity for symptomatic infections | 0.80, 0.90, **0.95**, 1.00 | - | - | 0.95 (0.9-0.99)[5] | (4) |
| Serology specificity | 0.990, 0.992, 0.994, 0.996, 0.998, **1.000** | - | - | 0.9984 (0.9911-100.0) | (5) |
| Yearly rate of seroreversion | 0.109 | - | - | 0.109 | (3) |

| | | | | | |
|---|---|---|---|---|---|
| after asymptomatic infection | | | | | |
| Yearly rate of seroreversion after symptomatic infection | 0.109 | - | - | 0.109 | (3) |

[1] Bold = Reference value fixed when exploring other parameter values, unless otherwise specified.

[2] Calculated as the reduction in the proportion of infected individuals that are symptomatic (cough, fever or loss of taste or smell) between those never previously positive and those positive <4 months ago.

[3] Assuming $VE_{sym}$ = 72.3% and $VE_{sym}$ = 66.5% is accurate for AstraZeneca and Janssen respectively, combinations of $VE_{in}$ and $VE_{pr}$ are calculated with Equation 1.

[4] $Minimum\ VE_{pr} = 1 - {}^{1}/_{p_s}$

[5] Elecsys Anti-SARS-CoV-2 assay (Roche, Basel, Switzerland).

***Supplementary Table 3. Rank regression uncertainty analysis for observed vaccine efficacy against infection for Oxford-AstraZeneca ChAdOx1.***

| | Estimate | Standard Error | T value | P value |
|---|---|---|---|---|
| **Intercept** | -14291.08 | 367.83 | -38.85 | <0.001 |
| **Vaccine efficacy against infection [1] (%)** | 0.66 | 0.00 | 173.41 | <0.001 |
| **Yearly hazard of infection (%)** | 0.52 | 0.03 | 19.98 | <0.001 |
| **Proportion symptomatic (%)** | 0.05 | 0.01 | 5.65 | <0.001 |
| **RR infection with prior infection (%)** | -0.01 | 0.03 | -0.35 | 0.725 |
| **RR symptoms with prior infection (%)** | -0.04 | 0.03 | -1.57 | 0.116 |
| **PCR test specificity (%)** | 143.02 | 3.68 | 38.86 | <0.001 |
| **PCR test sensitivity (asymptomatic) (%)** | 0.12 | 0.26 | 0.47 | 0.639 |
| **Days between asymptomatic PCR tests** | 0.85 | 0.03 | 26.10 | <0.001 |

[1] We assumed that the trial reported value for vaccine efficacy against symptomatic infection ($VE_{sym}$) was accurate, so as vaccine efficacy against infection increases, vaccine efficacy against progression decreases, such that $VE_{sym}$ is constant.

***Supplementary Table 4. Rank regression uncertainty analysis for observed vaccine efficacy against asymptomatic infection for Oxford-AstraZeneca ChAdOx1.***

| | Estimate | Standard Error | T value | P value |
|---|---|---|---|---|
| **Intercept** | -21990.99 | 958.88 | -22.93 | <0.001 |
| **Vaccine efficacy against infection [1] (%)** | 1.84 | 0.01 | 184.48 | <0.001 |
| **Yearly hazard of infection (%)** | 0.82 | 0.07 | 12.19 | <0.001 |
| **Proportion symptomatic (%)** | -0.72 | 0.02 | -31.87 | <0.001 |
| **RR infection with prior infection (%)** | -0.10 | 0.07 | -1.48 | 0.140 |
| **RR symptoms with prior infection (%)** | -0.07 | 0.07 | -1.09 | 0.275 |
| **PCR test specificity[1] (%)** | 218.80 | 9.59 | 22.81 | <0.001 |
| **PCR test sensitivity (asymptomatic) (%)** | -0.22 | 0.67 | -0.32 | 0.746 |
| **Days between asymptomatic PCR tests** | 0.48 | 0.09 | 5.63 | <0.001 |

[1] We assumed that the trial reported value for vaccine efficacy against symptomatic infection ($VE_{sym}$) was accurate, so as vaccine efficacy against infection increases, vaccine efficacy against progression decreases, such that $VE_{sym}$ is constant.

***Supplementary Table 5. Rank regression uncertainty analysis for observed vaccine efficacy against asymptomatic infection for Janssen Ad26.COV2.S.***

| | Estimate | Standard Error | T value | P value |
|---|---|---|---|---|
| **Intercept** | -2777.52 | 152.93 | -18.16 | <0.001 |
| **Vaccine efficacy against infection [1] (%)** | 2.05 | 0.02 | 101.15 | <0.001 |
| **Yearly hazard of infection (%)** | 0.37 | 0.14 | 2.73 | 0.006 |
| **Proportion symptomatic (%)** | -0.62 | 0.05 | -13.47 | <0.001 |
| **RR infection with prior infection (%)** | -0.22 | 0.14 | -1.59 | 0.113 |
| **RR symptoms with prior infection (%)** | -0.03 | 0.14 | -0.20 | 0.843 |
| **Serology test specificity (%)** | 26.72 | 1.52 | 17.53 | <0.001 |
| **Serology test sensitivity (asymptomatic infections) (%)** | 0.07 | 0.09 | 0.77 | 0.443 |
| **Serology test sensitivity (symptomatic infections) (%)** | -0.09 | 0.18 | -0.51 | 0.610 |

[1] We assumed that the trial reported value for vaccine efficacy against symptomatic infection ($VE_{sym}$) was accurate, so as vaccine efficacy against infection increases, vaccine efficacy against progression decreases, such that $VE_{sym}$ is constant.

### Supplementary Figures

***Supplementary Figure 1. Mechanisms of vaccine efficacy (VE).***

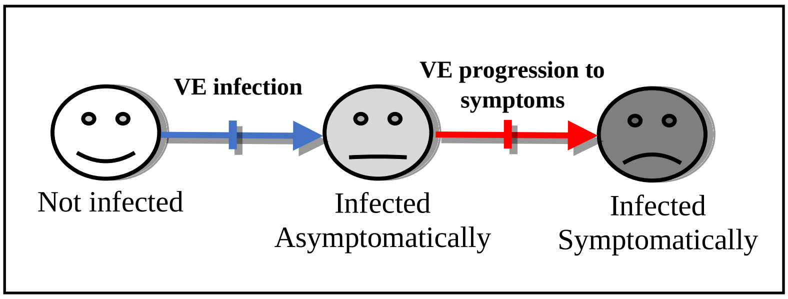

***Supplementary Figure 2. Flow diagram showing COVID-19 phase III vaccine trial model outline.*** The disease states are susceptible (unvaccinated) (S), vaccinated (V), infected asymptomatically ($I_A$), infected symptomatically ($I_S$), recovered from asymptomatic infection ($R_A$), recovered from symptomatic infection ($R_S$), waned antibodies after asymptomatic infection ($W_A$) and waned antibodies after symptomatic infection ($W_S$). Trial participants in $R_A$, $R_S$, $W_A$ and $W_S$ have the same level of natural immunity. Participants are seropositive in $R_A$, $R_S$ compartments only.

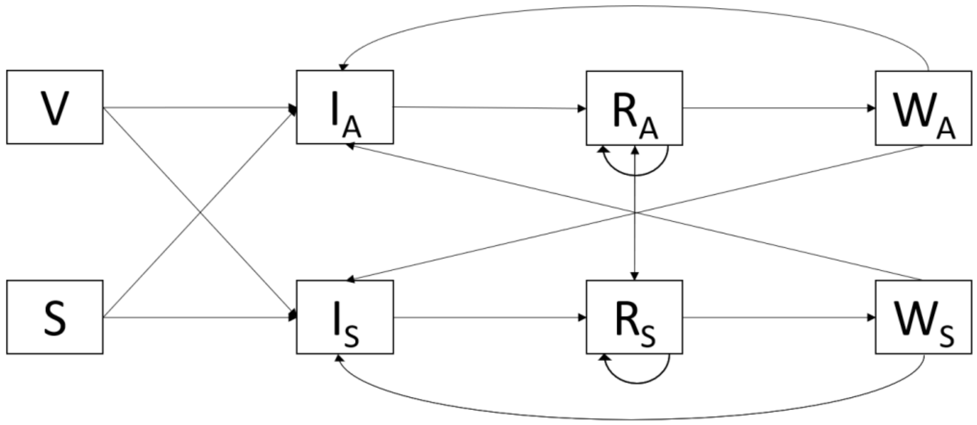

***Supplementary Figure 3. Delay distributions.*** A) Baseline time to infection, B) Time to seroconversion/recovery for first infection. Fit to data on the time to seroconversion from symptom onset from Long et al. (14), adding 5.5 days to each datapoint to account for the additional time from infection to symptom development, C) Time to seroreversion, D) Time to seroconversion/recovery for reinfection, after seroreversion.

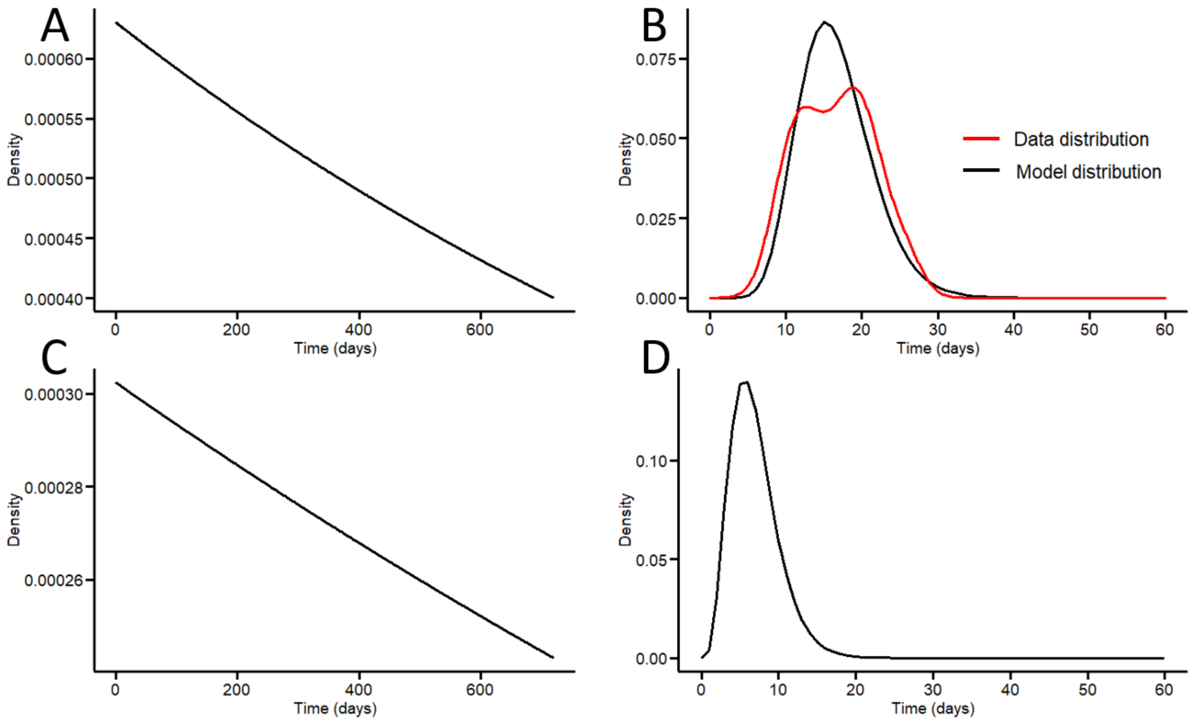

***Supplementary Figure 4. True vaccine efficacy against infection ($VE_{in}$) and vaccine efficacy against progression ($VE_{pr}$) as a function of true vaccine efficacy against asymptomatic infection ($VE_{asym}$) and true vaccine efficacy against symptomatic infection ($VE_{sym}$).*** Calculated using Equations 3 and 4, assuming the natural probability of developing symptoms upon infection is 0.67. E.g., for a vaccine with $VE_{sym}$= 70% and $VE_{asym}$= -20, $VE_{in}$ can be calculated to be 40% and $VE_{pr}$, 50%.

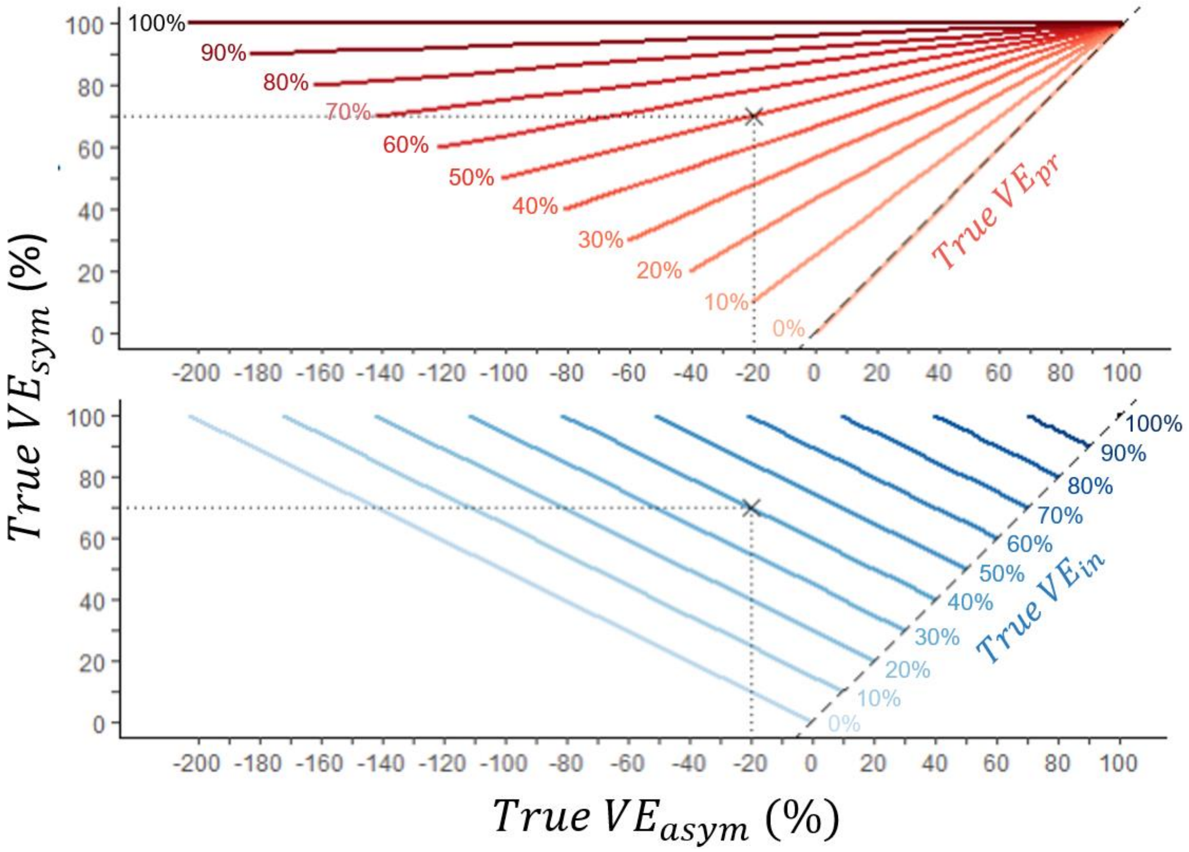

***Supplementary Figure 5. Change in estimated vaccine efficacy against symptomatic infection over a two-year follow-up, with efficacy calculated using 1-CIR.*** Time = 0 months represents 2 weeks post second dose. Sensitivity and specificity = 100%. True vaccine efficacies: vaccine efficacy against symptomatic infection $(VE_{sym}) = 70\%$, A) vaccine efficacy against infection $(VE_{in}) = 70\%$, vaccine efficacy against progression to symptoms $(VE_{pr}) = 0\%$, B) $VE_{in} = 50\%, VE_{pr} = 40\%$, C) $VE_{in} = 0\%, VE_{pr} = 70\%$. CIR = cumulative incidence ratio.

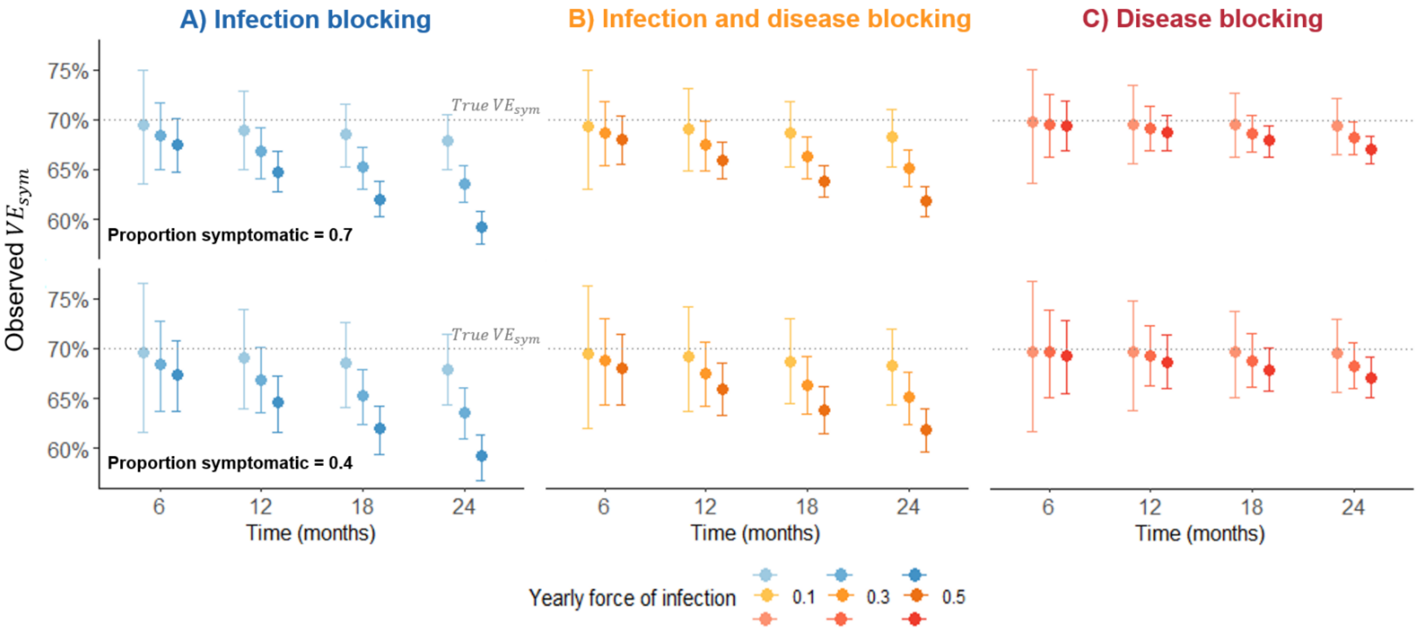

### References


1. Hall VJ, Foulkes S, Charlett A, Atti A, Monk EJ, Simmons R, et al. Do Antibody Positive Healthcare Workers Have Lower SARS-CoV-2 Infection Rates than Antibody Negative Healthcare Workers? Large Multi-Centre Prospective Cohort Study (The SIREN Study), England: June to November 2020. SSRN Electron J. 2021 Jan 15;2021.01.13.21249642.

2. Pritchard E, Matthews PC, Stoesser N, Eyre DW, Gethings O, Vihta KD, et al. Impact of vaccination on new SARS-CoV-2 infections in the United Kingdom. Nat Med. 2021;

3. Manisty C, Treibel TA, Jensen M, Semper A, Joy G, Gupta RK, et al. Time series analysis and mechanistic modelling of heterogeneity and sero-reversion in antibody responses to mild SARS-CoV-2 infection. EBioMedicine. 2021;65.

4. Harritshøj LH, Gybel-Brask M, Afzal S, Kamstrup PR, Jørgensen CS, Thomsen MK, et al. Comparison of 16 serological SARS-CoV-2 immunoassays in 16 clinical laboratories. J Clin Microbiol. 2021;59(5):2596–616.

5. Ainsworth M, Andersson M, Auckland K, Baillie JK, Barnes E, Beer S, et al. Performance characteristics of five immunoassays for SARS-CoV-2: a head-to-head benchmark comparison. Lancet Infect Dis. 2020;20(12):1390–400.

6. Voysey M, Costa Clemens SA, Madhi SA, Weckx LY, Folegatti PM, Aley PK, et al. Single-dose administration and the influence of the timing of the booster dose on immunogenicity and efficacy of ChAdOx1 nCoV-19 (AZD1222) vaccine: a pooled analysis of four randomised trials. Lancet. 2021;397(10277):881–91.

7. Skittrall JP, Wilson M, Smielewska AA, Parmar S, Fortune MD, Sparkes D, et al. Specificity and positive predictive value of SARS-CoV-2 nucleic acid amplification testing in a low-prevalence setting. Clin Microbiol Infect. 2021;27(3):469.e9-469.e15.

8. Emary KRW, Golubchik T, Aley PK, Ariani C V., Angus B, Bibi S, et al. Efficacy of ChAdOx1 nCoV-19 (AZD1222) vaccine against SARS-CoV-2 variant of concern 202012/01 (B.1.1.7): an exploratory analysis of a randomised controlled trial. Lancet. 2021;397(10282):1351–62.

9. Sadoff J, Gray G, Vandebosch A, Cárdenas V, Shukarev G, Grinsztejn B, et al. Safety and Efficacy of Single-Dose Ad26.COV2.S Vaccine against Covid-19. N Engl J Med. 2021 Apr 21;384(23):2187–201.

10. FDA. Vaccines and Related Biological Products Advisory Committee December 10, 2020 Meeting Briefing Document- FDA. 2020.

11. Hellewell J, Russell TW, Beale R, Kelly G, Houlihan C, Nastouli E, et al. Estimating the effectiveness of routine asymptomatic PCR testing at different frequencies for the detection of SARS-CoV-2 infections. BMC Med. 2021 Apr 27;19(1):106.

12. Oran DP, Topol EJ. The Proportion of SARS-CoV-2 Infections That Are Asymptomatic : A Systematic Review. Ann Intern Med. 2021 Jan 22;174(5):655–62.

13. Hall VJ, Foulkes S, Charlett A, Atti A, Monk EJM, Simmons R, et al. SARS-CoV-2 infection rates of antibody-positive compared with antibody-negative health-care workers in England: a large, multicentre, prospective cohort study (SIREN). Lancet. 2021;397(10283):1459–69.

14. Long QX, Liu BZ, Deng HJ, Wu GC, Deng K, Chen YK, et al. Antibody responses to SARS-CoV-2 in patients with COVID-19. Nat Med. 2020;26(6):845–8.
